# A Prospective Longitudinal Study of Health-Related Quality of Life and Psychological Wellbeing after an Implantable Cardioverter Defibrillator in Patients with Genetic Heart Diseases

**DOI:** 10.1101/2021.04.26.21256086

**Authors:** Lieke M. van den Heuvel, Tanya Sarina, Joanna Sweeting, Laura Yeates, Kezia Bates, Catherine Spinks, Catherine O’Donnell, Samuel F. Sears, Kevin McGeechan, Christopher Semsarian, Jodie Ingles

## Abstract

**Background:** Genetic heart diseases often affect young people, can be clinically heterogeneous and pose an increased risk of sudden cardiac death (SCD). The implantable cardioverter defibrillator (ICD) is a lifesaving therapy. Impacts on prospective and long-term psychological and health-related quality of life (HR-QoL) after ICD implant in patients with genetic heart diseases are unknown. We investigate the psychological functioning and HR-QoL over time in patients with genetic heart diseases who receive an ICD, and identify risk factors for poor psychological functioning and HR-QoL.

**Methods:** A longitudinal, prospective study design was used. Patients attending a specialised clinic and diagnosed with a genetic heart disease, for which they received an ICD between May 2012 and January 2015, were eligible. Baseline surveys were completed prior to ICD implantation with five-year follow-up after ICD implant. We measured psychological functioning (Hospital Anxiety Depression Scale, Florida Shock Anxiety Scale), HR-QoL (Short-Form 36v2) and device acceptance (Florida Patient Acceptance Scale).

**Results:** There were 40 patients with an inherited cardiomyopathy or arrhythmia syndrome included (mean age 46.3 ± 14.2 years; 65.0% males). Mean psychological and HR-QoL measures were within normative ranges during follow-up. After 12 months, 33.3% and 19.4% of participants showed clinically elevated levels of anxiety and depression, respectively. Longitudinal mixed effect analysis showed significant improvements from baseline to first follow-up for the overall cohort, with variability increasing after 36 months. Low education and female gender predicted worse mental HR-QoL and anxiety over time, while comorbidities predicted depression and worse physical HR-QoL.

**Conclusion:** While the majority of patients with a genetic heart disease adjust well to their ICD implant, a subset of patients’ experience poor psychological and HR-QoL outcomes.

## INTRODUCTION

Genetic heart diseases include inherited cardiomyopathies and arrhythmia syndromes. They are characterized by clinical heterogeneity, with outcomes ranging from minimal symptoms to severe heart failure and sudden cardiac death (SCD) [1]. For those considered at increased risk of SCD, due to factors such as family history, unexplained syncope, prior cardiac arrest or sustained ventricular tachycardia, an implantable cardioverter defibrillator (ICD) is recommended [2]. While potentially life-saving, ICD implantation carries the risk of inappropriate shocks and complications in patients with genetic heart diseases [3].

ICD implantation can have a detrimental impact on psychological functioning and health-related quality of life (HR-QoL) in a subset of patients [4-6]. Several factors may contribute to patients being vulnerable to negative psychological outcomes and worse HR-QoL, such as female gender, type D personality (i.e., a combination of negative affectivity and social inhibition), increased anxiety and depression at baseline, ICD shocks, comorbidities and a young age at implant (<45 years) [7-12]. Due to heterogeneity in study methodologies, existing literature shows mixed evidence, with multiple studies reporting similar or even improved HR-QoL after ICD implantation compared to standard medical therapy [13-16] or pacemakers [17].

Research to understand psychological outcomes and HR-QoL has been conducted mostly in the setting of coronary artery disease [4,6-8,10-13,16,17], where patients are generally older, i.e., mean age >60 years. Comparatively fewer studies have focused on patients with genetic heart diseases. While most patients show good adaptation [18-23], there is an important subgroup that report symptoms of anxiety, depression and posttraumatic stress symptoms [18,19,22]. Most studies have so far been cross-sectional, making it difficult to draw conclusions regarding causation. We performed a prospective, longitudinal cohort study with five-year follow-up, which aimed to investigate the psychological functioning and HR-QoL over time in patients diagnosed with genetic heart diseases who receive an ICD. Further, we investigated risk factors for poor psychological functioning and HR-QoL.

## METHODS

### Participants

Patients attending a specialised multidisciplinary genetic heart disease clinic in Sydney, Australia who underwent implantation of an ICD between May 2012 and January 2015 were invited to participate. Individuals were eligible for the study if they were aged 18 years or older, had English-skills sufficient to complete the survey, and had received a diagnosis of an inherited arrhythmia syndrome or inherited cardiomyopathy. Patients considered likely to be recommended an ICD were approached prior to their initial clinic appointment and invited to complete a general survey focused on their psychological wellbeing and quality of life. Some patients were approached in hospital prior to surgical implantation of their ICD. Only patients who eventually underwent ICD insertion and completed at least one follow-up survey were considered study participants. The study was approved by the local institutional ethics committee and all patients provided written informed consent.

### Data collection

Clinical and demographic data were obtained from the Australian Genetic Heart Disease Registry, and/or the medical record, which are continually updated and reviewed by cardiologists and other health professionals. Data collected included basic demographics, including gender, age, ethnicity, education level (low=below university, high=university or higher) and socioeconomic status (SES, based on the Index of Relative Socioeconomic Advantage and Disadvantage (IRSAD)). The IRSAD is used by the Australian Bureau of Statistics to summarize information about the economic and social conditions of people living within certain areas (24). Clinical information, such as clinical diagnosis, presence of comorbidities, and family history were collected. Details regarding ICD implantation and outcomes, including number of shocks at follow-up, were also collected.

### Patient surveys

Several validated scales were administered at baseline and several follow-up time points, 1-3 months, 6 months and every 12 months post-ICD implant. Participants were contacted by phone or email and invited to complete the follow-up survey. Scales included:

#### The Medical Outcomes Short Form 36 version 2 (SF-36v2)

The Medical Outcomes Short Form 36 version 2 (SF-36v2) is a validated scale measuring HR-QoL [25,26]. The SF-36v2 is comprised of 36 items and provides a score (range 1-100) for eight sub-domains (physical functioning, role limitations due to physical health, general health, social functioning, bodily pain, vitality, role limitations due to emotional health and mental health) and two composite scores (physical component score; PCS and mental component score; MCS). Only PCS and MCS scores were included in this analysis. MCS and PCS scores were converted to Australian weighted T-scores. The weighted T-scores range from 0 (worst possible health) to 100 (best possible health) [27]. A score of 50 is the mean score for the general Australian population.

#### Hospital Anxiety and Depression Scale

The Hospital Anxiety and Depression Scale (HADS) is a validated measure of psychological wellbeing over the last seven days, used extensively in the hospital setting [28]. It is comprised of 14 items from which summary scores of anxiety and depression can be determined (range 0 to 21). A cut-off score of ≥8 is used to describe clinically elevated levels of anxiety and depression that may warrant further investigation, as previously shown in an HCM population [29].

#### Florida Patient Acceptance Scale & Florida Shock Anxiety Scale

The Florida Patient Acceptance Scale (FPAS) and the Florida Shock Anxiety Scale (FSAS) were included [30,31]. The FPAS comprises 18 items and is used to assess acceptance of the ICD. It provides measures of return to life, positive appraisal, device-related distress and body image concerns [30]. The FSAS includes 10 items and aims to determine the patient’s anxiety related to the consequences and triggers of an ICD shock [31]. Both the FPAS and FSAS use a 5-point Likert scale and provide a summary score of overall acceptance and anxiety, respectively. A higher score on the FPAS indicates higher acceptance of the device, whereas a higher score on the FSAS indicates a higher level of anxiety regarding shocks.

### Statistical analysis

Data were analysed using SAS Studio statistical software (Version 5.2) and R Studio (Version 1.2.1335). Data were visualised using the R ggplot2 package. Sample characteristics and survey-responses are described as means (SD) or median (IQR), as appropriate. The longitudinal changes in HADS, SF-36v2, FSAS and FPAS were estimated using linear mixed models with a random intercept (lme4 package, version 1.1-21), which assumes that missing data are missing at random [17]. Repeated measurements were nested within subjects. Time was included as a categorical variable in the analyses. Due to the extensive follow-up period of five years, many other factors might have influenced our psychological and HR-QoL outcomes. Therefore, baseline measures of age category (i.e., young <40 years old and older ≥40 years old), gender, education level, and the presence of comorbidities and shocks were included as fixed effects in the mixed model analysis. The p-values are shown for these analyses, however due to known limitations of their use with the lmer4 package, confidence intervals have been primarily used in interpretation of results.

## RESULTS

### Population characteristics and response rates

In total, 91 participants were approached prior to ICD implant, where there was a suspicion an ICD may eventually be recommended. Of these, 63 (69.2%) completed a baseline survey, including 23 (36.5%) who completed the baseline survey prior to discussion with the doctor about an ICD recommendation, 38 (60.3%) after an ICD was recommended and 1 (1.6%) which was uncertain. Overall, 42 (66.7%) went on to have an ICD, with 40 going on to complete at least one follow-up survey (considered study participants), and 2 declining to complete further surveys. There were 4/63 (6.3%) who declined an ICD and were deemed ineligible, and 17/63 (27.0%) where an ICD was deemed not indicated.

Table 1 shows the demographic and clinical characteristics of the 40 participants at baseline. Mean age was 46.3 ±4.0 years (range 19.8 to 66.1 years), 26 (65.0%) were male and 33 were of European ethnicity (83%). A third (14/40, 35.0%) had a university education and most participants had a high socioeconomic status (23/40, 57.5%). The most common genetic heart disease was HCM (30/40, 75.0%) and 43.0% of patients (17/40) had comorbidities, including stroke, kidney disease, diabetes, cancer, asthma and arthritis. Only five patients (13.0%) had shocks during follow-up (range: 1-3 shocks).

**TABLE 1:** Sociodemographic and clinical characteristics at baseline.

| **Characteristics** | **N (%)** |
| --- | --- |
| **Total (N)** | 40 |
| **Male gender** | 26 (65.0) |
| **Age range (mean, SD)** | 19.8 - 66.1 (46.3, 14.2) |
| **European ethnicity** | 33 (82.5) |
| **Education level\*** |  |
| Low | 26 (65.0) |
| High | 14 (35.0) |
| **Socioeconomic status at baseline\*\*** |  |
| Low | 4 (10) |
| Moderately low | 4 (10) |
| Moderately high | 9 (22.5) |
| High | 23 (57.5) |
| **Disease type** |  |
| HCM | 30 (75) |
| DCM | 1 (2.5) |
| ACM | 2 (5.0) |
| BrS | 4 (10) |
| LQTS | 3 (7.5) |
| **Comorbidities present** | 17 (42.5) |
| **Patients with shocks during follow-up** | 5 (12.5) |
| Range shocks per patient | 1-3 |

Abbreviations: HCM, Hypertrophic Cardiomyopathy; DCM, Dilated Cardiomyopathy; ACM, Arrhythmogenic Cardiomyopathy; BrS, Brugada Syndrome; LQTS, Long QT Syndrome.
\*Education level was defined as low (below university) and high (university or higher)
\*\*Socioeconomic status was defined by the Australian IRSAD index (24). Data are presented in categories: low = percentiles 0-24; moderately low = 25-49; moderately high = 50-74; high = 75-100)

### Impact on psychological functioning

Figure 1 shows the predicted values of the psychological and HR-QoL outcome measures. Mean HADS-anxiety and depression scores were below the cut-off score of 8 at baseline and each follow-up time point, although large standard deviations were observed (see Table 2). Almost half of participants (19/40, 47.5%) indicated increased anxiety (HADS anxiety ≥8) at baseline and 33.3% (12/36) beyond 12 months post-implant. Overall, 14 participants (35%) scored ≥8 at one point >12 months during follow-up. Of those patients who had clinically elevated levels of anxiety at baseline, 14/19 (42.8%) also showed clinically elevated levels during follow-up.

**FIGURE 1:**
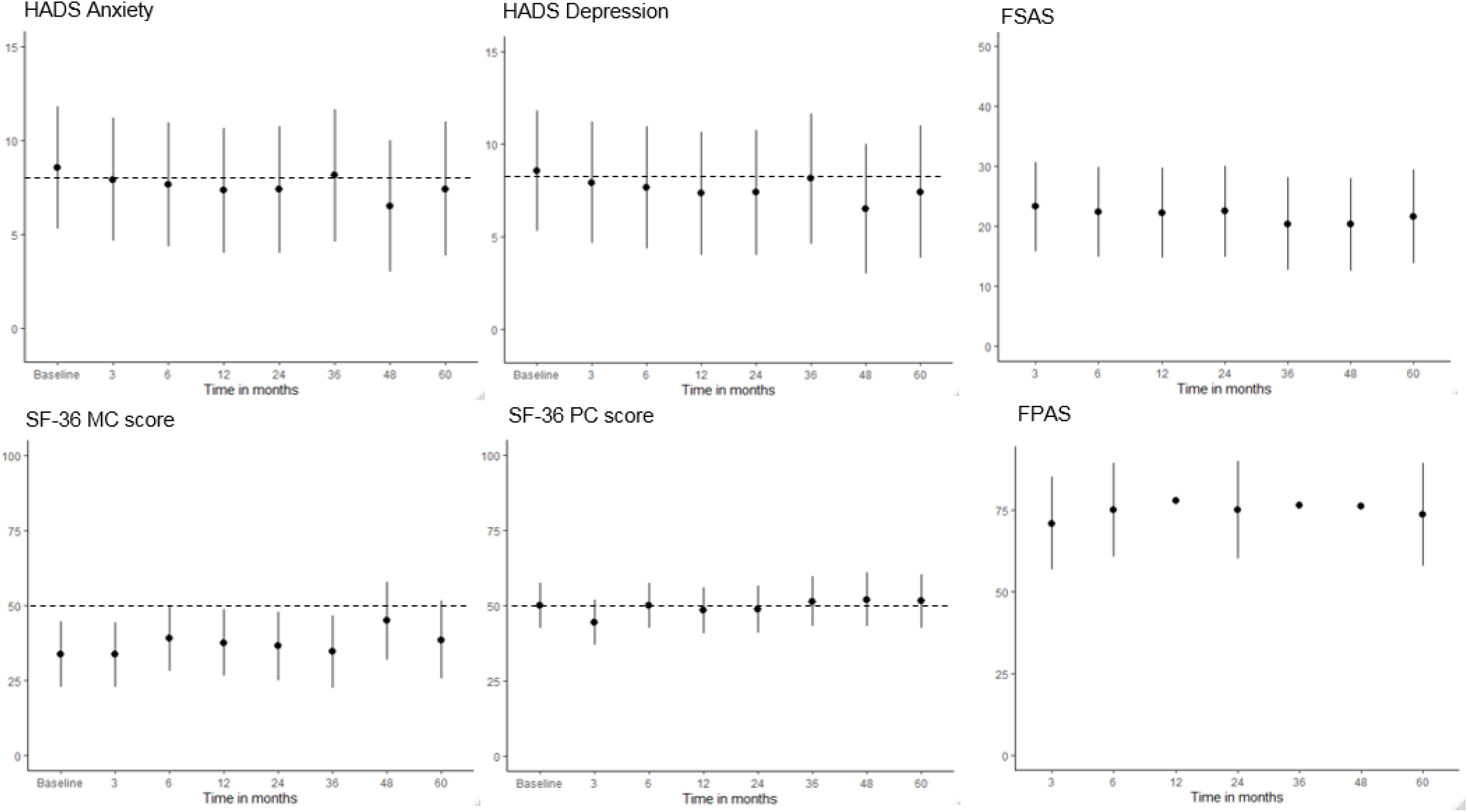
Predicted values of outcome measures with 95% confidence intervals over follow-up based on mixed model analysis. The dotted line indicates the cut-off score and z-score for respectively HADS and SF-36 scores. Abbreviations: *HADS, Hospital Anxiety and Depression Scale (HADS); FSAS, Florida Shock Anxiety Scale; MC, Mental Component; PC, Physical Component*.

**TABLE 2:**
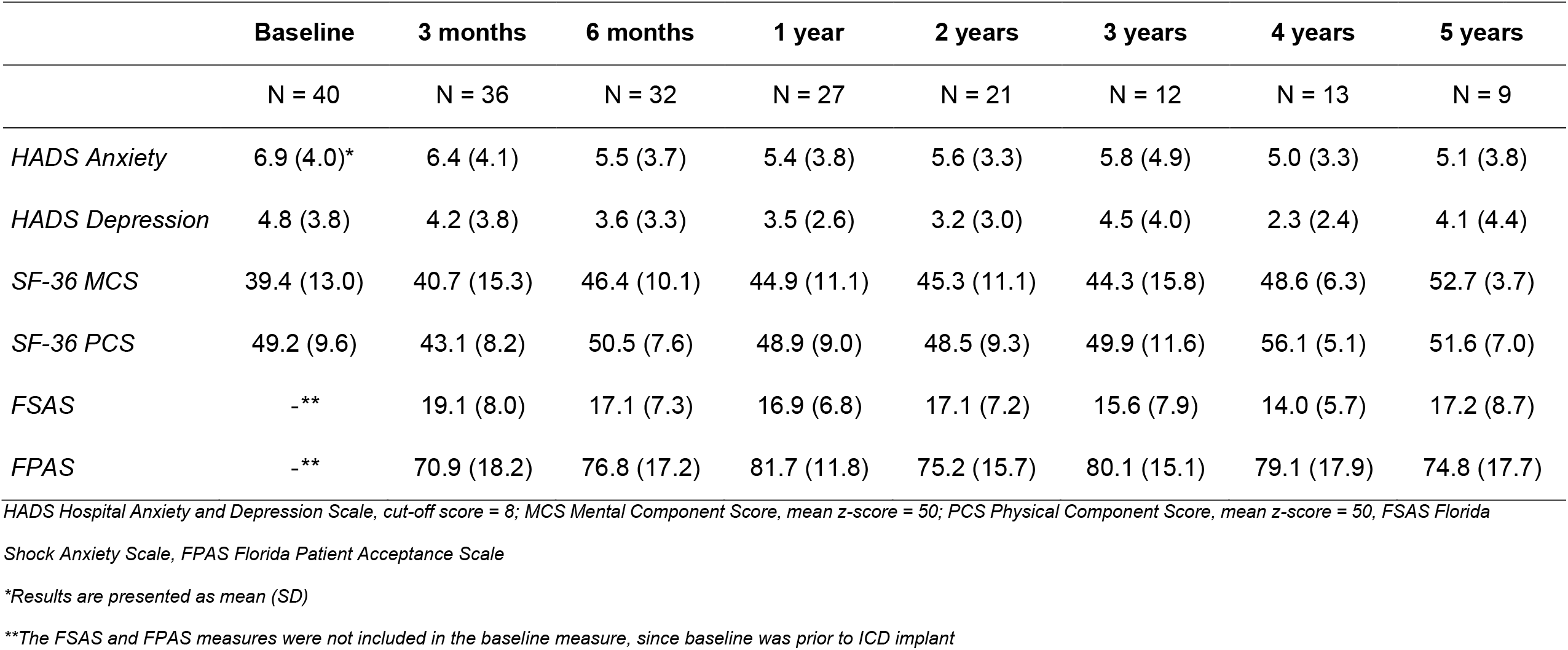
Psychological and health-related quality of life outcome measures per timepoint.

Anxiety symptoms initially increased but gradually improved over time, compared to baseline, with a significant improvement in HADS anxiety score after 4 years (95% CI -14.05--2.87, p=0.017). In addition, participants with university-level education (HADS anxiety score 2.55 points lower, 95% CI -4.98--0.11, p=0.070) and male participants (HADS anxiety score 2.47 points lower, 95% CI -4.96 – 0.00, p=0.084) had less anxiety symptoms. Predictors including age, ethnicity and the presence of comorbidities or shocks, did not show an effect. Further, none of these predictors showed a significant interaction with time.

With respect to depression, 25.0% (10/40) reported symptoms of depression (HADS depression ≥8) at baseline, and 19.4% (7/36) at 1-year post-implant. Overall, 5 participants reported depression symptoms at one point >1 year during follow-up. At follow-up, HADS-depression scores showed an improvement over time compared to baseline. Participants with comorbidities were predicted to have worse depression symptoms (HADS depression score 2.17 points higher, 95% CI 0.04-4.27, p=0.081) than participants without comorbidities (see Table 3).

**TABLE 3:** Estimated fixed effects of predictors for anxiety and depression.

| Measure | Variable | Category | Mean change | 95% CI |  | P for main effect<sup>a</sup> | P for interaction effect with time<sup>b</sup> |
| --- | --- | --- | --- | --- | --- | --- | --- |
|  |  |  |  | Lower | Upper |  |  |
| *HADS anxiety* | Time | Baseline | *Reference* |  |  |  | *NA* |
|  |  | 3 months | 0.63 | -1.91 | 3.18 | 0.685 |  |
|  |  | 6 months | -1.42 | -4.15 | 1.26 | 0.395 |  |
|  |  | 1 year | -1.83 | -4.99 | 1.30 | 0.345 |  |
|  |  | 2 years | -1.20 | -5.45 | 3.12 | 0.650 |  |
|  |  | 3 years | 1.65 | -3.46 | 6.91 | 0.604 |  |
|  |  | **4 years** | **-8.26** | **-14.05** | **-2.87** | 0.017 |  |
|  |  | **5 years** | **-0.00** | **-5.47** | **-5.64** | 0.999 |  |
|  | Age | ≥ 40 | **-0.07** | **-2.46** | **-2.41** | 0.962 | 0.077 |
|  | Gender | Male | -2.47 | -4.96 | 0.00 | 0.084 | 0.917 |
|  | Education | **University** | **-2.55** | **-4.98** | **-0.11** | 0.070 | 0.387 |
|  | Ethnicity | White | 0.57 | -2.66 | 3.77 | 0.756 | 0.772 |
|  | Comorbidities | Present | 1.21 | -1.37 | 3.77 | 0.409 | 0.409 |
|  | Shocks | Yes | -0.32 | -3.82 | 3.19 | 0.873 | 0.324 |
| *HADS depression* | Time | Baseline | *Reference* |  |  |  | *NA* |
|  |  | 3 months | 0.94 | -1.87 | 3.75 | 0.587 |  |
|  |  | 6 months | -0.11 | -3.08 | 2.89 | 0.952 |  |
|  |  | 1 year | -1.56 | -5.00 | 1.91 | 0.461 |  |
|  |  | 2 years | -1.02 | -5.63 | 3.77 | 0.724 |  |
|  |  | 3 years | 2.25 | -3.28 | 8.06 | 0.516 |  |
|  |  | 4 years | -2.45 | -8.67 | 3.53 | 0.511 |  |
|  |  | 5 years | -3.49 | -9.42 | 2.84 | 0.350 |  |
|  | Age | ≥ 40 | -0.96 | -3.00 | 1.18 | 0.430 | 0.408 |
|  | Gender | Male | -1.40 | -3.45 | 0.63 | 0.241 | 0.399 |
|  | Education | University | -1.96 | -3.96 | 0.04 | 0.095 | 0.916 |
|  | Ethnicity | White | 2.64 | -0.05 | 5.27 | 0.091 | 0.808 |
|  | Comorbidities | **Present** | **2.17** | **0.04** | **4.27** | 0.081 | 0.755 |
|  | Shocks | Yes | 1.81 | -1.07 | 4.69 | 0.281 | 0.503 |
| *SF-36 Mental Component Score (MCS)* | Time | Baseline | *Reference* |  |  |  | *NA* |
|  |  | 3 months | 1.29 | -7.06 | 9.53 | 0.793 |  |
|  |  | 6 months | 8.81 | 0.20 | 17.30 | 0.083 |  |
|  |  | 1 year | 7.03 | -3.17 | 17.19 | 0.243 |  |
|  |  | **2 years** | **15.13** | **3.27** | **26.97** | 0.032 |  |
|  |  | 3 years | -6.49 | -21.62 | 8.33 | 0.461 |  |
|  |  | 4 years | 12.48 | -2.24 | 27.18 | 0.153 |  |
|  |  | 5 years | 7.97 | -14.44 | 30.00 | 0.543 |  |
|  | Age | ≥ 40 | 0.08 | -8.59 | 8.42 | 0.986 | 0.455 |
|  | Gender | **Male** | **8.79** | **0.34** | **17.18** | 0.072 | 0.222 |
|  | Education | University | 2.81 | -5.29 | 10.85 | 0.544 | 0.227 |
|  | Ethnicity | White | 3.38 | -5.21 | 12.14 | 0.489 | *NA<sup>c</sup>* |
|  | Comorbidities | Present | 2.19 | -6.44 | 10.83 | 0.658 | 0.532 |
|  | Shocks | Yes | -7.80 | -17.12 | 1.57 | 0.142 | *NA<sup>c</sup>* |
| *SF-36 Physical Component Score (PCS)* | Time | Baseline | *Reference* |  |  |  | NA |
|  |  | **3 months** | **-8.65** | **-14.46** | **-2.85** | 0.013 |  |
|  |  | 6 months | 2.72 | -3.25 | 8.71 | 0.443 |  |
|  |  | 1 year | -0.55 | -7.64 | 6.63 | 0.897 |  |
|  |  | 2 years | -1.61 | -9.87 | 6.75 | 0.744 |  |
|  |  | 3 years | 5.58 | -4.85 | 16.06 | 0.368 |  |
|  |  | 4 years | -4.19 | -14.53 | 6.08 | 0.492 |  |
|  |  | 5 years | 6.08 | -9.48 | 21.55 | 0.508 |  |
|  | Age | ≥ 40 | -1.65 | -8.78 | 4.22 | 0.637 | 0.787 |
|  | Gender | Male | -0.22 | -6.18 | 6.07 | 0.951 | 0.783 |
|  | Education | University | 3.65 | -2.19 | 9.49 | 0.275 | 0.118 |
|  | Ethnicity | White | 2.82 | -3.25 | 9.72 | 0.428 | NA<sup>c</sup> |
|  | Comorbidities | **Present** | **-8.99** | **-15.14** | **-2.61** | 0.014 | 0.573 |
|  | Shocks | Yes | 5.43 | -1.45 | 12.36 | 0.162 | NA<sup>c</sup> |
| *Florida Shock Anxiety Scale (FSAS)* | Time | 3 months | *Reference* |  |  |  | NA |
|  |  | 6 months | 0.60 | -3.21 | 4.49 | 0.809 |  |
|  |  | 1 year | -2.04 | -6.57 | 2.52 | 0.483 |  |
|  |  | 2 years | 0.45 | -5.65 | 6.66 | 0.909 |  |
|  |  | 3 years | 2.59 | -4.65 | 10.13 | 0.582 |  |
|  |  | **4 years** | **-11.77** | **-19.76** | **-3.87** | 0.023 |  |
|  |  | **5 years** | **8.15** | **0.43** | **16.12** | 0.107 |  |
|  | Age | ≥ 40 | -4.16 | -8.28 | 0.51 | 0.109 | 0.121 |
|  | Gender | Male | -1.62 | -6.69 | 3.17 | 0.566 | 0.304 |
|  | Education | University | -3.97 | -8.75 | 0.94 | 0.155 | 0.922 |
|  | Ethnicity | White | -0.91 | -7.27 | 5.09 | 0.796 | 0.374 |
|  | Comorbidities | Present | 0.49 | -4.52 | 5.46 | 0.864 | 0.628 |
|  | Shocks | Yes | 2.59 | -4.23 | 9.59 | 0.511 | 0.212 |
| *Florida Patient Acceptance Scale (FPAS)* | Time | 3 months | *Reference* |  |  |  | NA |
|  |  | **6 months** | **16.48** | **4.81** | **27.89** | 0.028 |  |
|  |  | **1 year** | **21.97** | **8.13** | **35.34** | 0.013 |  |
|  |  | 2 years | -1.87 | -21.20 | 15.79 | 0.873 |  |
|  |  | 3 years | 2.74 | -19.84 | 24.21 | 0.845 |  |
|  |  | **4 years** | **43.77** | **20.01** | **67.64** | 0.005 |  |
|  |  | 5 years | 6.36 | -17.80 | 29.31 | 0.672 |  |
|  | Age | ≥ 40 | -1.54 | -12.91 | 8.86 | 0.810 | 0.200 |
|  | Gender | Male | 9.23 | -2.37 | 20.99 | 0.178 | 0.323 |
|  | Education | University | 4.22 | -7.27 | 15.61 | 0.526 | 0.330 |
|  | Ethnicity | White | 5.21 | -8.93 | 19.93 | 0.532 | 0.066 |
|  | Comorbidities | Present | -2.59 | -14.22 | 9.17 | 0.702 | 0.094 |
|  | Shocks | Yes | -14.32 | -30.51 | 2.00 | 0.132 | 0.369 |

Reference categories are: gender = female, education level = no university, ethnicity = white ethnicity, age category = young age, shocks = no shocks, comorbidities = no comorbidities present. Abbreviations: HADS = Hospital anxiety and depression scale.
<sup>a</sup>Based on Wald's t-test
<sup>b</sup>Based on type III ANOVA table with Sattthertwaite's method
<sup>c</sup>Model including interaction between timepoint and shocks and ethnicity respectively is rank deficient. The interactions timepoint \*shocks and timepoint\*ethnicity were therefore excluded from the model.

### Impact on HR-QoL

The mean mental component score (MCS=39.4) was overall lower (worse) than the physical component score (PCS=49.2) and both gradually improved over time, although large standard deviations were observed (Table 2).

Mental component scores gradually improved over time (see Table 4), with significant improvement after two years (95% CI 3.27 -26.97, p=0.032). Predictors including age, education, ethnicity and the presence of comorbidities or shocks, did not significantly contribute to the model. However, male patients scored 8.79 points better than female patients on mental HR-QoL (95% CI 0.34-17.18, p=0.072). Also, none of the predictors included in the model showed a significant interaction with time.

Over time a gradual improvement in physical component scores was observed, after an initial significant decrease at 3 months follow-up (95% CI -14.46--2.85, p=0.013). The presence of comorbidities showed a significant effect on predicted physical component score values, with participants with comorbidities scoring 8.99 points lower than participants without comorbidities (95% CI -14.14--2.61, p=0.014). We observed no significant interaction effects with time.

### Shock anxiety

Mean scores for shock anxiety gradually increased over time, though the sample size decreased substantially with time (Table 2). A significant improvement in shock anxiety was observed at 4 years (95% CI -19.76--3.87, p=0.023), compared to the first measure (1-3 months; Table 3). The predictors included in the model did not show a significant effect nor significant interactions with time.

### Device acceptance

Mean scores on the FPAS scale show a gradual improvement in patient acceptance of their ICD after the first 6 months, with large standard deviations (Table 2). FPAS scores improved over time, with a significant improvement after 6 months (95% CI 4.81 – 27.89, p=0.028), one year (95% CI 8.13 – 35.34, p=0.013) and after 4 years (95% CI 20.01 – 67.64, p = 0.005), compared to the first three months after ICD implantation. Predictors did not show a significant contribution to the model, and none of the predictors showed significant interactions with time.

## DISCUSSION

Despite being life-saving, implantation of an ICD can have important psychological functioning and health-related quality of life impacts. With most evidence based on older cardiovascular disease populations, in contrast patients diagnosed with a genetic heart disease are often younger in age at diagnosis, have minimal or no symptoms, and must deal with the heritable nature of the disease [18,19,32]. We show that while mean values of psychological and HR-QoL measures were within the normal range over time, the large variability in confidence intervals and standard deviations highlighted the wide range of responses. Furthermore, a sizeable subgroup of patients showed clinically elevated levels of anxiety and depression during early follow-up. Lower education level and female gender were identified as a predictor for worse mental HR-QoL and anxiety, while the presence of comorbidities predicted symptoms of depression and worse physical HR-QoL.

Our findings support prior cross-sectional studies in the genetic heart disease population, which show that while most patients adjust well to their ICD, an important subgroup do have ongoing psychological difficulties [18,19,21-23]. Due to our longitudinal study design, we show for the first time that overall psychological wellbeing and mental HR-QoL improve over time. However, it is important to note the increasing variability for all measures observed after three years follow-up. Longitudinal studies investigating the effect of ICD implant in a heterogeneous group of patients report a gradual improvement in outcomes over time [33-35], or indicate an initial improvement [14]. However, follow-up periods described in these studies differ substantially, and as we found, it is challenging to control for the numerous other life events that occur with such long-term follow-up periods, which impact on emotional wellbeing and HR-QoL.

Higher general anxiety scores in ICD patients over time were observed in female patients and patients with lower education levels. Female gender has previously been identified as a predictor for poor psychological functioning and HR-QoL [7,12]. Previous work examining ICD implantation in individuals with coronary artery disease found no association between education level and worse psychological functioning [37,38]. Wong et al. [39] found an association between low education level and worse depression scores, but reported no association with anxiety. It is important to note however, that many studies have not included education level in their analyses [18-22]. While previous research has identified a younger age as a risk factor for poor psychological outcomes and HR-QoL in patients with genetic heart disease [19,21,22], this was not observed in our longitudinal data. Type D personality and being optimistic have been identified as strong predictors for psychological outcomes in studies in ICD patients with coronary artery disease and myocardial infarction [11,38]. Furthermore, the perception of social support has been considered a predictor for psychological adjustment to an ICD and may therefore be an interesting predictor as well [37]. These factors have not been included in research on long-term psychological outcomes after ICD implantation in patients with genetic heart disease so far [18,19,22].

Our study design has several limitations, including the relatively small sample size and response rate drop off over time. Collecting baseline surveys prior to ICD implantation presented a major challenge in recruitment for this study and led to our small sample size. This may have limited our power to identify relevant predictors of poor psychological functioning of HR-QoL. Furthermore, we cannot be sure if drop-out over time influenced our outcomes. In addition, very few patients in our cohort received shocks, meaning prospective evaluation of the impact of shocks could not be reliably assessed. Previous research has identified the presence and number of shocks as an important predictor of poor psychological outcomes, and therefore our data may underestimate the effect of ICD implantation on psychological functioning and HR-QoL. Of note, a large Swedish study of ICD patients (N>3000) suggested that concern about a potential shock, rather than the shock itself, predicted poor outcomes [12].

Overall, our findings indicate that although a majority of patients adjust well to their ICD, those with poor baseline psychological functioning and HR-QoL pre-implantation, females, lower education level and presence of comorbidities, are at increased risk of experiencing anxiety, depression and/or worse HR-QoL post-implant. Ideally, these patients should be identified in clinic and monitored carefully during follow-up. Psychosocial support and interventions might be effective in diminishing distress and reducing anxiety and depressive symptoms in these patients. While psychological interventions, including psycho-education and cognitive behavioural therapy, have shown to be effective in ICD patients with other, non-genetic heart diseases [40], no intervention research has been performed in those with genetic heart diseases specifically. Due to the unique circumstances that these patients face, including the young age at diagnosis, often being relatively asymptomatic despite having high risk of SCD, and the heritable nature of disease, tailored interventions are likely to be more effective. Since our findings suggest that there is increasing variability after three years, support programs and interventions should ideally incorporate a longer-term follow-up for patients who are at risk of poor psychological functioning.

## CONCLUSIONS

We report the first longitudinal self-report survey study to evaluate psychological functioning and health-related quality of life after ICD implantation in patients with genetic heart diseases. We show normative psychological outcomes over time, although the large variability observed highlights the diverse responses. An important subgroup of patients showed clinically elevated levels of anxiety and depression during follow-up, with low education level, female gender and the presence of comorbidities being predictors of poor psychological functioning and HR-QoL. Patients vulnerable to developing poor psychological outcomes should ideally be identified in clinic and carefully monitored. Tailored psychosocial support and interventions might be effective to diminish distress and relieve anxiety and depressive symptoms in this unique patient group.

## Data Availability

Data is available from the corresponding author at reasonable request and following consultation with our local ethics committee.

